# The natural history of homonymous hemianopia revisited

**DOI:** 10.1101/2022.10.06.22280668

**Authors:** Elizabeth L. Saionz, Matthew R. Cavanaugh, Brent A. Johnson, Donald Harrington, Geoffrey K. Aguirre, Krystel R. Huxlin

## Abstract

**Objective:** To re-evaluate the longitudinal progression of stroke-induced homonymous visual field defects using strictly automated perimetry (Zeiss Humphrey Systems), rigorous inclusion/exclusion criteria, and quantitative analyses.

**Methods:** A retrospective chart review of stroke patients diagnosed with “homonymous hemianopia”, who underwent monocular Humphrey visual field (HVF) perimetry using the 24-2 SITA standard pattern from 2011-2019, was conducted at a large US academic medical center. Reliable tests (<20% fixation losses, false positives, and false negatives) were identified and analyzed with generalized estimating equations to extract temporal trends in perimetric mean deviation (PMD) and deficit area.

**Results:** Of 532 patients with “homonymous hemianopia”, sequential, reliable HVFs were only available for 36 patients in the right eye, and 30 patients in the left eye, ranging from 7 days to 58 months post-stroke. Both PMD and deficit area improved early, within the first 3 months post-stroke; however, this was followed by a subsequent decline in performance >1 year post-stroke. Changes were similar between eyes.

**Conclusion:** We discovered that a large portion of occipital stroke patients do not receive comprehensive ophthalmologic follow-up and, even then, only a fraction of HVFs performed are reliable enough for rigorous analysis. Nonetheless, reliable HVFs in such patients confirmed early visual improvement after stroke, consistent with prior reports. However, in contrast with prior, qualitative reports, there was no stability of the deficit beyond 6 months post-stroke; instead, gradual worsening erased the initial spontaneous improvement, especially >1 year post-stroke.

## Introduction

Homonymous hemianopia (HH) is a major cause of morbidity after stroke.^1^ Clinically, it is assessed by visual perimetry, with Humphrey or other automated perimetry considered most sensitive and reliable.^1-6^ Advantages of automated perimetry include good test-retest fidelity and validity metrics, including active fixation monitoring and measurement of false positive and negative rates. Yet only one group has investigated the natural history of stroke-related HH with this tool.^7^ Their study expanded upon prior findings from confrontation assessment^8-10^ and Goldmann perimetry,^11-13^ using binary logistics to show that patients with HH were likely to experience spontaneous visual field improvement in the first 3-6 months post-stroke, with no improvement beyond 6 months, concluding that deficits hereafter become stable and permanent.^7^ However, this study included non-stroke participants, assessments other than Humphrey perimetry, did not consider visual field reliability metrics, and used a binary model permitting only “improvement” or “non-improvement” which was later interpreted as stability.^7^

Recent work has both confirmed and challenged aspects of this early model for hemianopic progression. In a small study of chronic HH (>6 months post-stroke), the usual care group showed worsening of Humphrey visual fields (HVFs) over time.^14^ Although humans are known to experience trans-synaptic retrograde degeneration of early visual pathways after occipital strokes,^15-25^ testing the hypothesis that this is associated with slowly degrading vision requires a non-binary analysis of visual defects. Here, we conducted a retrospective review of reliable HVFs in occipital stroke patients, evaluating HVF change longitudinally, within subject, to revisit and parametrize the progression of HH defects.

## Methods

### Selection of patients

Medical records were retrospectively screened for all patients with a diagnosis of “homonymous hemianopia” or related term (ICD-9-CM 368.46 and ICD-10-CM H53.46) seen by the department of ophthalmology at a large US academic medical center (the University of Rochester) from 2011-2019. Patients captured by this screening search were evaluated for inclusion according to whether they were assigned a procedure code for visual field testing (CPT 92083). If they received visual field testing, their medical records were then reviewed for exclusion criteria including visual field defect etiology not due to stroke (e.g., brain tumor, traumatic brain injury, multiple sclerosis), additional neurologic or ophthalmologic diagnoses that could affect visual fields (e.g., glaucoma, diabetic retinopathy, macular degeneration, optic neuropathy, advanced cataracts, and other significant vascular and neurological diseases affecting the retina, ocular lenses and ocular media). Finally, for included patients, etiology of stroke (embolic-ischemic or hemorrhagic) was noted in each case, if available, from visit notes or radiological reports. This study was approved by the University of Rochester Medical Center Research Subjects Review Board.

### Visual field testing

After patients were screened for inclusion, raw data files for all of their available automated perimetric visual fields were collected (Humphrey Field Analyzer II-i750, Zeiss Humphrey Systems, Carl Zeiss Meditec, Atlanta, GA). Only fields using the Humphrey 24-2 test pattern with SITA Standard algorithm were then assessed for meeting reliability criteria, defined as having the eye-tracker “on”, reliability check “on”, and with less than 20% each of fixation losses, false positives, and false negatives.^14^ For inclusion in the final analysis, patients had to have multiple visits with reliable HVFs in at least one eye and they could not participate in a visual training research study or clinical trial.

Each reliable monocular visual field was analyzed for perimetric mean deviation (PMD), a Humphrey-derived metric (Zeiss Humphrey Systems) that compares the patient’s hill of vision to that of an age-corrected normal population. We also interpolated visual sensitivity at the 55 test locations (including the fovea) of each monocular field using a natural-neighbors function to a resolution of 0.1 deg, as previously described.^14^ This interpolated field was then used to calculate the deficit area (DA), defined as the size of the region of the HVF where luminance sensitivity was <10 dB, consistent with the United States Social Security Administration’s criteria for legal blindness.^26^ The primary outcome measure for the present study was change in the PMD; a secondary endpoint was change in DA.

### Statistical methods

We used ordinary descriptive statistics to summarize demographic variables and outcomes. As mentioned above, the principal outcome was within-patient change in PMD. For each patient in the cohort, the change in PMD was calculated as the difference between the first-available monocular field PMD and every subsequent field PMD for that eye; fields collected on the day of stroke (day 0) or >10,000 days post-stroke were excluded. Because of the observational nature of the study, and because patients did not always have reliable tests in both eyes at each visit, the data set included a variable number of fields meeting inclusion criteria for each eye of each patient throughout the evaluation period.

To best capture the trajectory of change in HVFs, we restricted our analysis to patients whose first visual field test was performed within the first 90 days post-stroke, then assessed PMD and deficit area changes in four time-windows, according to the time of subsequent visual field tests (1-90 days, 91-180 days, 181-360 days, and >360 days post-stroke). While we investigated using more granular windows of time from 30 days and upwards, this produced a significant loss of precision in estimation resulting from smaller numbers of patients per window; ultimately, 90-day windows proved to be most valid. We modeled the change in PMD/deficit area via generalized estimating equations (GEE),^27^ as these yield valid statistical inferences for repeated measures data through a working correlation matrix. Binary indicators for the four time-windows (1-90 days, 91-180 days, 181-360 days, and >360 days) served as independent predictors in the statistical regression model. The window for 1-90 days was set as the model intercept, with each time-window thereafter modeled as change from the intercept.

Eye (right/OD or left/OS) was also included as an independent, binary predictor in the model. Robust standard error (SE) estimates, corresponding test statistics, and p-values were reported using an exchangeable working correlation model assumption. Calculations were computed in the R statistical software, version 4.1.1 (R Core Team, cran.r-project.org); data are available upon reasonable request.

## Results

### Characteristics of patients and visual fields

A preliminary screening of medical records identified many patients who could potentially be captured by our study; however, after screening for inclusion and reliability criteria, only a small fraction remained eligible. Of 532 patients diagnosed with HH or equivalent (see Methods) between 2011 and 2019, only 410 underwent visual field testing **(figure 1, A**). Of these patients, 192 were excluded from further analysis here for clinical reasons (108 for a neurologic diagnosis additional to occipital stroke, and 84 for an ophthalmologic diagnosis). Of the remaining 218 patients, 25 had only Goldmann visual field testing done, leaving 193 patients with HVFs. However, most of these patients (53%) received only a single visual field exam, and thus, could not contribute to the present analysis (**figure 1, B**). Of those remaining (**figure 2, A**), multiple, reliable 24-2 HVFs were performed in 74 unique patients in the right eye (OD; 204 HVFs) and 62 unique patients in the left eye (OS; 177 HVFs). These visual fields were collected at times ranging from 0 to 2,987 days post-stroke (**figure 2, B and C**); one patient was an outlier, with two visual fields in each eye collected 10,745 and 10,992 days post-stroke, causing them to be removed from subsequent analyses. Remaining patients had from 2-7 reliable visual field visits per person, for both OD and OS.

**Figure 1.**
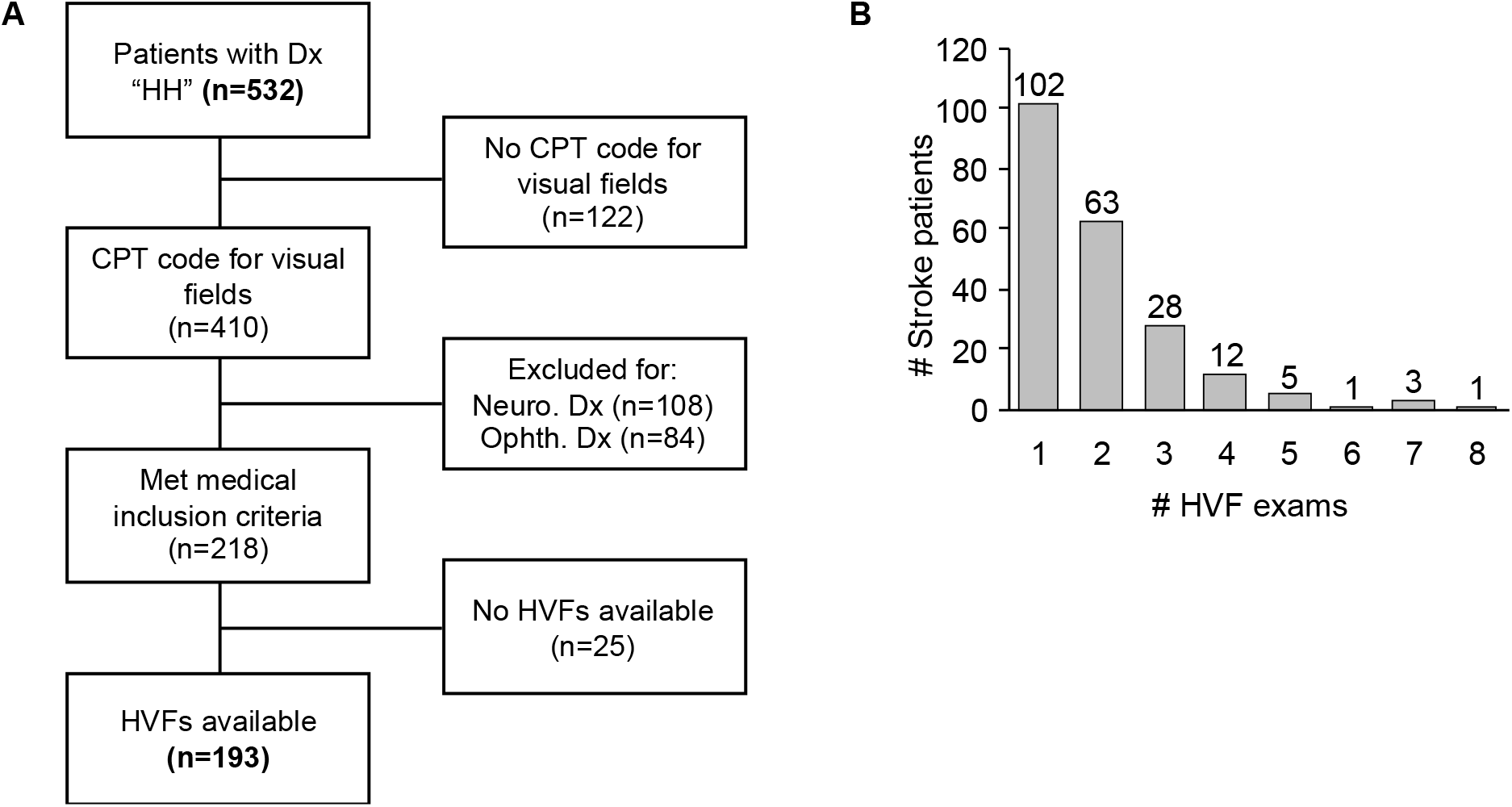
Selection of patients for analysis (A) Flow diagram of patient selection process with inclusion/exclusion criteria. HH: homonymous hemianopia. (B) Histogram of number of Humphrey visual field (HVF) examinations per patient. Over 50% of patients had only a single exam.

**Figure 2.**
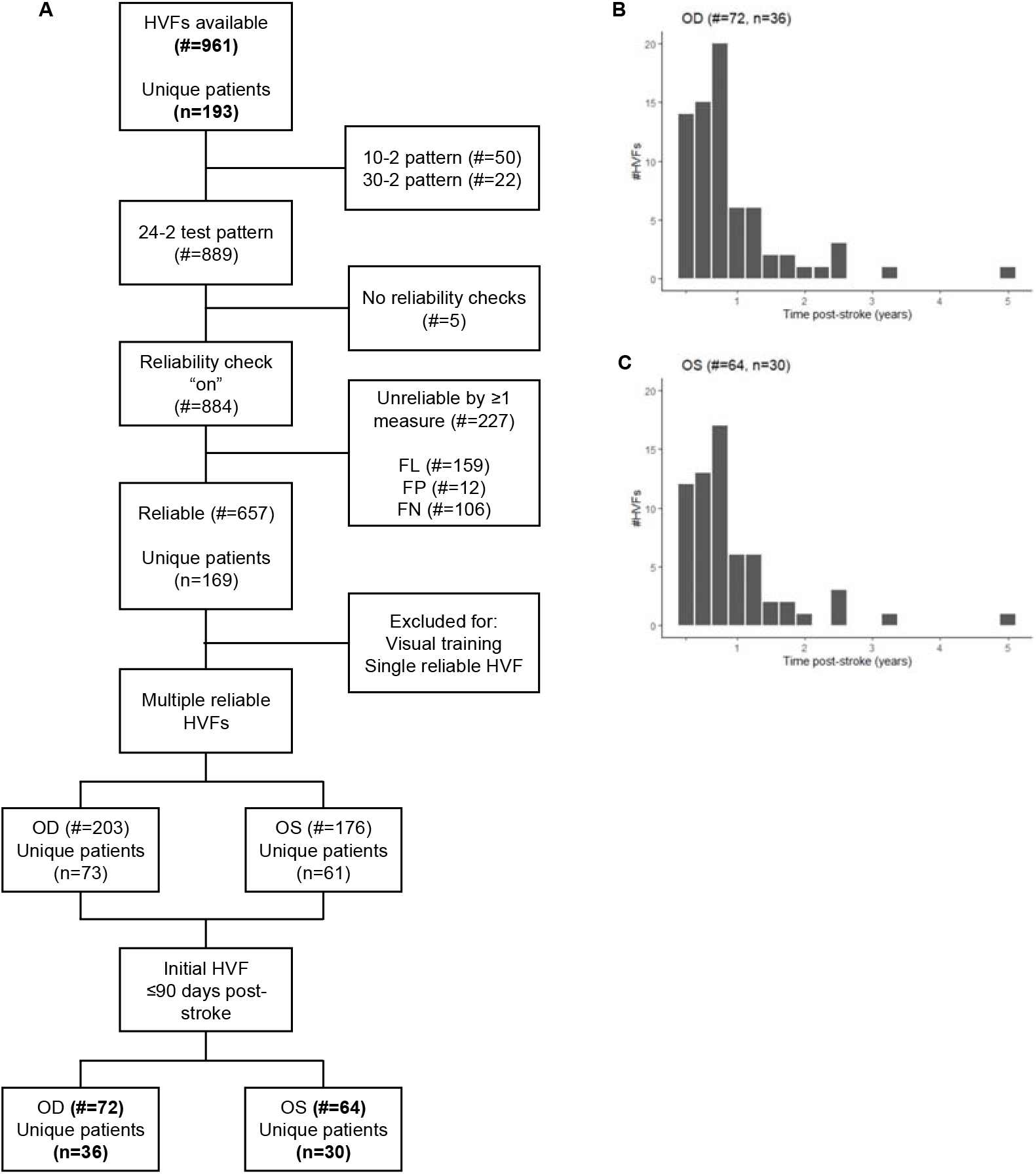
Selection of visual field tests for analysis (A) Flow diagram of HVF selection per HVF reliability criteria. (B) Histogram of HVF of the right eye (OD) included in final analysis (excluding one outlier with two HVFs) by time post-stroke. (C) Histogram of HVFs of the left eye (OS) included in final analysis (excluding one outlier with two HVFs) by time post-stroke. HVF: Humphrey visual field, FL: fixation loss, FP: false positive, FN: false negative, OD: right eye, OS: left eye, #: number of HVFs, n: number of subjects.

The highest rate of visual field testing was in the first 3 months post-stroke (29% OD, 26% OS), and less than half of the tests (37% OD; 40% OS) were from more than a year after the stroke. Finally, most strokes were embolic/ischemic in origin, with 133 OD/114 OS; there were 42 OD/37 OS strokes identified as hemorrhagic, and an additional 29 OD/26 OS unknown in origin.

In sum, multiple, reliable HVFs were obtained in only a fraction of occipital stroke patients with HH treated in a 9-year period between 2011 and 2019 at a large US academic medical center.

### GEE model of change in PMD over time after stroke

From this set of repeat reliable HVFs, we first assessed the change in PMD over time, using the difference between first visual field collected within 90 days of stroke and subsequent visual fields. More reliable field pairs were available for OD (n=36, with 72 observations) than OS (n=30, with 64 observations) (**figure 2, A**). They ranged from a first visual field test at 2-90 days post-stroke and subsequent visual field tests from 16 to 1726 (OD) and 7 to 1726 (OS) days.

Boxplots illustrating changes in PMD from group to group are displayed in **figure 3, A** and the estimates from the marginal model regression fits via GEE are given in **Table 1**. The model suggests that PMD initially increased (i.e. improved) within the first 90 days post-stroke, then remained about the same until approximately 360 days post-stroke, after which time PMD declined (i.e. worsened). From the regression fits, PMD increased by 2.06 dB (robust standard error 0.55) from the baseline assessment to another assessment within 90 days post-stroke. From 90 to 360 days post-stroke, the change in PMD from baseline was not statistically different from change within the first 90 days post-stroke. However, the change in PMD from baseline to assessments beyond 360 days was statistically significant at -0.87 dB (robust standard error 0.44), suggesting a consistent reduction in global visual field sensitivity during this time window. On average, despite significant improvement in the first 90 days post-stroke, the final PMD was only just above the initial PMD. There was no significant difference between eyes.

**Figure 3.**
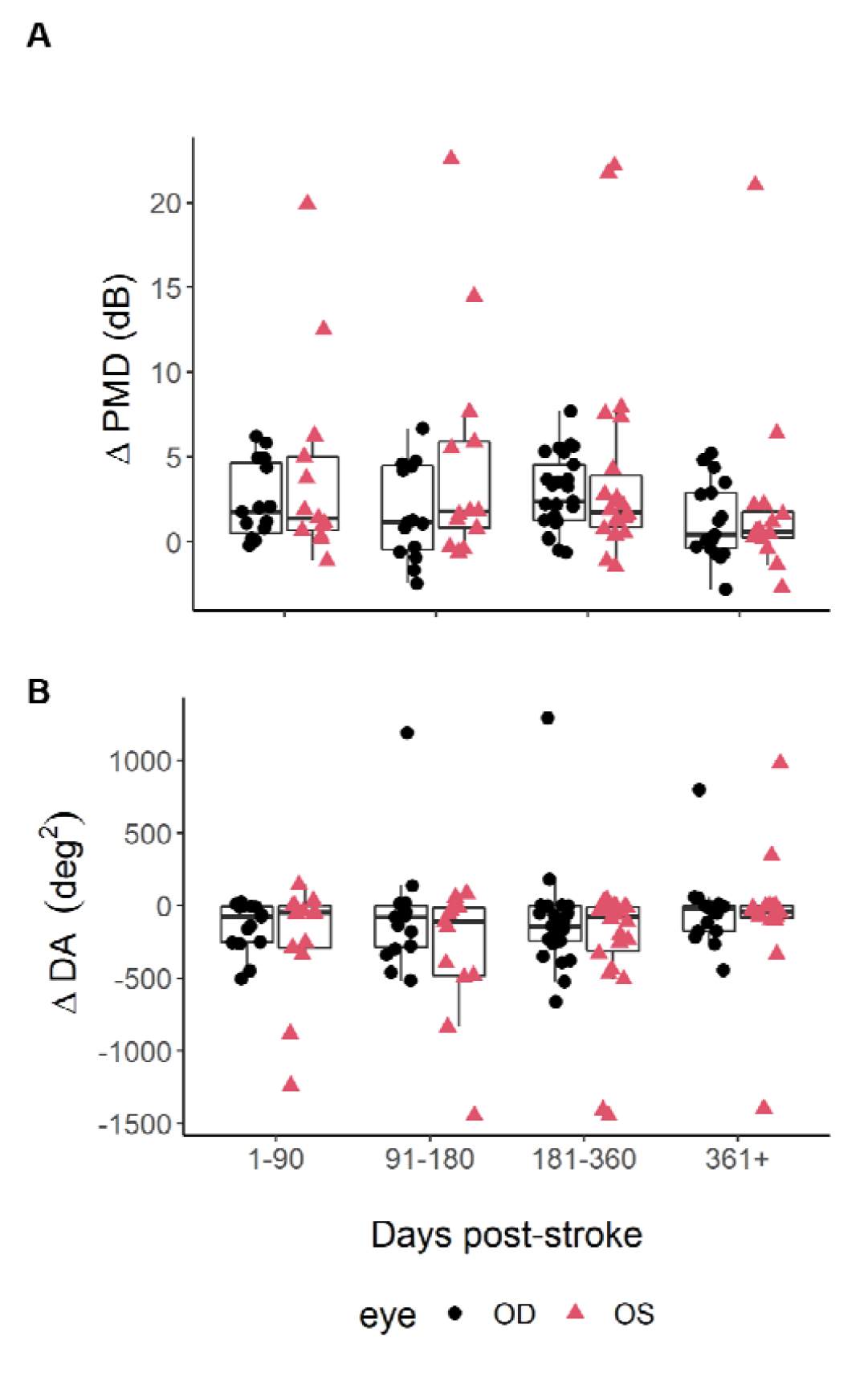
Boxplots of the within-patient changes in (A) perimetric mean deviation (PMD), and (B) deficit area (DA) from the first available HVF post-stroke to every subsequent HVF in 1-90, 91-180, 181-360, 361-1,800 days post-stroke. Boxes indicate median (horizontal line), interquartile range (box), and range (vertical lines). Analyses are restricted to patients whose first available field was within 90 days of their stroke date.

**Table 1.** Model of change in outcome (perimetric mean deviation (PMD) and deficit area (DA)) from first visit post-stroke to each subsequent visit, restricted to patients whose first visit post-stroke was within 90 days of their stroke date. Coefficient and robust standard error (RSE) estimates are derived from generalized estimating equations (GEE) with exchangeable working correlation model. The model intercept below is interpreted as the average difference between the initial visit 1-90 days post-stroke and the first subsequent visit post-stroke; remaining rows are interpreted as an average difference of differences: e.g. the average difference between differences 1-90 days post-stroke (from first visit) and differences 91-180 days post-stroke (from first visit). Data from both eyes were analyzed together and modeled as a separate term in the statistical model, but the average difference between the left vs. right eye over time (effect “eye”) was not statistically significant at the nominal rate.

| | ΔPMD | | ΔDA | |
| --- | --- | --- | --- | --- |
| Effects | Estimate (RSE) | p-value | Estimate (RSE) | p-value |
| 1-90 days (intercept) | **2.06 (0.55)** | **<0.01\*\*** | **-97 (52)** | 0.06 |
| 91-180 days | -0.70 (0.58) | 0.23 | 21 (36) | 0.56 |
| 181-360 days | 0.22 (0.58) | 0.71 | -1.3 (29) | 0.96 |
| 361+ days | **-0.87 (0.44)** | **0.049\*** | **68 (31)** | **0.03\*** |
| Eye | 1.06 (1.42) | 0.45 | -87 (99) | 0.38 |

### GEE model of change in deficit area (DA) over time after stroke

Because the PMD is a global measure of visual sensitivity that encompasses both intact and impaired regions of the central visual field, it was key to ascertain whether its temporal dynamics were driven primarily by changes associated with the visual defect or by global changes in sensitivity across the entire visual field. To address this, we separately analyzed changes in DA over time, finding them to be consistent with changes in PMD. The DA model suggested an initial decrease (i.e. improvement) in the area of the HVF containing sensitivities <10 dB within the first 90 days post-stroke, followed by a relative plateau until approximately 360 days post-stroke, after which time DA grew again and returned to approximately the initial deficit size (**Figure 3, B** and **Table 1)**. On average, the deficit area shrank by 97 deg^2^ (robust standard error 52) in the first 90 days after stroke; it then remained essentially unchanged until 360 days post-stroke, but then expanded by 68 deg^2^ (robust standard error 31) beyond 360 days post-stroke. Because the total area of the 24-2 HVF is constant (1,545 deg^2^), the present findings with respect to DA suggest that the major factor driving the change in PMD is indeed a fluctuation in the size of the area of the deficit. Again, there was no significant difference between eyes.

## Discussion

Based on a few studies of patients with mixed etiology, a variety of tools and non-quantitative assessments, the natural history of homonymous visual field defects has long been assumed to follow a simple trajectory, consisting of spontaneous improvement in the first 3-6 months post-stroke, with deficits becoming stable and permanent thereafter. Here, we re-examined the natural history of such visual field defects after restricting our analysis to visual cortex strokes, a single instrument/approach to measure these defects, and strict inclusion/exclusion criteria for both the test results and the patients. In a departure from previous studies, we relied solely on automated (Humphrey) perimetric visual field measurements with clear, validated standards for test reliability. We evaluated a relatively uniform cohort of stroke-only patients, using well-defined, continuous metrics of visual sensitivity change across time. The GEE method was chosen to account properly for the correlation among repeated observations, i.e. multiple differences in PMD or deficit area per patient. The parameter estimates in Table 1 are said to be consistent even when the assumed working correlation model is incorrect; our parameter estimates, and findings, are robust in this statistical sense. In addition, we modeled change in PMD from baseline (set at 1-90 days post-stroke) using 90-day windows of time and removed those patients who did not have a baseline PMD. We investigated finer windows of time, e.g. 30- and 60-day windows, but the trends were not statistically significant due to the loss of precision in estimation resulting from the smaller number of patients per window. Parametric nonlinear models of time would be a natural remedy for this statistical problem for a future study in which dense PMD measurements are collected.

Our GEE analysis showed that visual field sensitivity improved in the first 3 months post-stroke, then stayed essentially stable through the remainder of the first year, before regressing to around baseline levels, at least within the timeframe examined (up to 58 months post-stroke). As such, these findings confirm aspects of prior studies showing early spontaneous improvement.^7, 10, 12, 13^ However, they also challenge the dogma that chronic post-stroke visual field sensitivity improves to a stable and *permanent* level. Instead, we now report that sensitivity begins to decline, in a way that becomes consistent and significant, starting at 1 year post-stroke.

These changes were first evident in terms of PMD, a global measure of sensitivity across the entire visual field. However, the fact that they were also evident in terms of deficit area across the study population suggests that the driving element for these trends was blind field visual performance rather than visual sensitivity across the entire central visual field. For disease states like HH in which vision loss is restricted to discrete regions of the visual field, deficit area ultimately may be a more accurate measurement of disease progression compared to PMD. Still, the PMD has the added advantage of being an age-adjusted metric; as such, the fact that the trends in PMD for later times post-stroke (>1 year) concurred with those in deficit area suggest that both were related to the underlying pathophysiology of HH and not solely due to an age-related decline in vision.

The present findings of a late decline in visual sensitivity in the blind field are consistent with prior work from our group,^14^ which examined changes in HVF sensitivity in 5 patients with chronic HH (>180 days post-stroke); in this cohort, first HVFs were collected 444 days post-stroke (standard deviation 147 days), and second HVFs were performed 42 to 399 days later.^14^ Although the sample size was very small, the data showed evidence of worsening in visual fields in the absence of a training intervention. Visual sensitivity declined by >6 dB over an area approximately 8.7 deg^2^ (standard deviation 4.5 deg^2^) of the binocular 24-2 HVF, equating to a 0.06 dB (standard deviation 0.14 dB) loss of PMD across the 5 participants. This surprising initial observation prompted the present study. It remains true that an even larger sample size than we were able to analyze here may be necessary for us to estimate the decline in blind field visual performance with adequate precision.

Our original intent in the present study was indeed to capture a large sample size, on the order of several thousand visual fields. However, we encountered several major roadblocks that prevented us from achieving this goal. These can be grouped broadly into: 1) lack of repeat automated perimetry in occipital stroke patients, and 2) poor quality of automated perimetry tests performed.

Automated (Humphrey) perimetry is the clinical gold standard for measuring visual field loss in HH.^6^ The prevalence of visual field loss in the *general* stroke population may be as high as 30%,^1^ and repeat perimetry is important for appropriate diagnosis as well as for tracking progression.^6, 28-30^ Nevertheless, some stroke patients are unaware of their own visual field deficit, ^5^ and many stroke centers do not routinely test for vision loss. ^2^ In the present study, out of >500 patients identified with HH at a large academic center, only ∼75% underwent HVF testing over a nine-year period after their stroke, and less than half of this subset of patients further received repeat testing. Indeed, we may never accurately estimate the total number of patients with HH who were not evaluated by an ophthalmologist or neuro-ophthalmologist. We posit that patients fail to receive appropriate follow-up testing because physicians may believe that there is little benefit to formal visual fields if no validated rehabilitative interventions are available.^1^ Moreover, medicine is steeped in the dogmatic belief that stroke-induced visual field loss is stable and permanent once patients reach 6 months post-stroke. This assumption has been reinforced by observations since Holmes first described the condition in 1918,^7, 9-13^ but few studies used any form of perimetry at all; only Zhang and colleagues^7^ used automated perimetry, the acknowledged clinical gold standard. But even in that study, multiple non-equivalent forms of perimetry were utilized, no reliability metrics were applied to the HVFs, no quantitative assessment of defect size was performed, and a mixed patient population - including vision loss not due to occipital stroke - was used.

Even when automated perimetry is ordered, the exam must be administered reliably in order to have utility. Our study focused on the Humphrey 24-2 test pattern, which is the most common test ordered for stroke patients with suspected visual field loss.^6^ We initially attempted a similar analysis at a second medical center (the Hospital of the University of Pennsylvania) but were unable to complete it, as none of the 10,000 HVFs screened were collected using the “reliability check” during testing. Even at our primary study site - where technicians were rigorously trained in performing perimetry and instructed to follow a standard script for every exam - only 74% of the HVFs analyzed met our reliability criteria. The most common reason for test failure was excessive fixation losses (18% of HVFs). Many factors contribute to fixation instability in HH, including difficulty following test instructions, stroke-induced inability to control eye movements, and abnormal fixation behavior.^31, 32^ Yet, fixational stability is critical for HVF test reliability, as inappropriate fixation could alter the spatial extent and severity of the visual field defect, or even obscure it entirely. Additionally, prior work showed that excessive false positive rates during testing were the HVF reliability criterion with greatest impact on PMD.^33^ This was also meaningful here because false positives may present a particular problem for patients with HH experiencing visual hallucinations, which tend to be more common early after stroke. However, in our sample, only 1% of HVFs were rejected due to excessive false positives. False negative responses were the third reliability criterion considered here, which can cause an underestimate of visual field sensitivity. They should be easily addressed through proper instruction by the administering technician, but in our sample, 12% of HVFs had excessive false negatives. Finally, we should note that less than half of patients with reliable HVFs who met our medical inclusion criteria had more than one reliable test in at least one eye. Reliability was slightly higher for right eyes, which we posit could occur because they are usually tested first. As such, patient fatigue may have contributed to poorer test quality for left eyes. Permitting patients longer breaks between eye exams could improve reliability of second eye tests. Overall, our findings suggest that 1) standardizing technician training protocols, and 2) implementing quality control across institutions treating this patient population, would together augment understanding of the progression of HH post-stroke, and improve our ability to rigorously evaluate the impact of therapeutic interventions.

Aside from implementing strict reliability criteria, another major advantage of our analysis over prior studies was our use of GEE to evaluate PMD and deficit area shifts over time, while controlling for individual differences in the natural history population such as disease severity, timing of HVFs, and number of HVFs. The GEE analysis showed that, across patients, PMD initially increased, and deficit area correspondingly decreased, in the first 3 months after stroke; these values remained stable throughout the rest of the first year, and then subsequently regressed. Such a pattern is suggestive of initial resolution of cytotoxic edema and inflammation, followed by the development and increasing impact of trans-synaptic retrograde degeneration.^15-25^ In trans-synaptic retrograde degeneration, the loss of targets in primary visual cortex leads sequentially to the death of relay neurons in the ipsilesional dorsal lateral geniculate nucleus of the thalamus, and then to the loss of retinal ganglion cells in the affected portion of both eyes.^15-25^ Our results are thus consistent with the temporality of trans-synaptic retrograde degeneration: experiments in non-human primates have found histological evidence of degenerative changes as early as 100 days after occipital lesioning,^18^ and optical coherence tomography in humans has found thinning of the retinal nerve fiber layer in the affected retinal region as early as one month post-stroke.^20^ Loss of these early visual sensory substrates could constrain the effectiveness of visual restoration treatments^34, 35^;a recent study of vision training therapy after stroke found that training-induced recovery of visual fields in the chronic post-stroke period was indeed limited by shrinkage of the optic tracts.^25^ Together with the present work, these results underscore the necessity of investigating the underpinnings, precise chronology, and individual variability of vision loss after visual cortex strokes to facilitate the development and timely implementation of targeted therapeutics.

In conclusion, the present study highlighted a surprising lack of reliable documentation of visual field defects in patients with stroke-induced HH with automated perimetry in a large US academic medical center. Restricting the original sample size to only those patients with HVFs that met rigorous reliability and inclusion criteria, we then applied quantifiable metrics and robust statistical modeling. Our findings confirmed that early visual fields are indeed likely to improve within three months post-stroke. However, they challenged the conventional wisdom that defects then stabilize in the chronic period. Instead, we found chronic visual fields to worsen over time, a possible functional consequence of progressing retrograde degeneration of early visual pathways. These results further support the notion that perimetry for HH serves three important purposes: diagnostic accuracy, monitoring, and functional assessment.^6, 36^ However, this population remains underserved. With approximately a quarter million new patients per year suffering from cortical stroke-induced vision loss in the United States alone,^1^ our findings underscore the need to rigorously monitor deficit progression in order both to better manage patients safety and functionality in everyday life, and to better target and assess the impact of therapeutic interventions for this condition. Our clinical standards and practice for HH need to support our patients and those endeavoring to treat them.

## Data Availability

All data produced in the present study are available upon reasonable request to the authors.

